# Performance Assessment of ECG Delineators on Single-Lead Wearable Ambulatory Data

**DOI:** 10.64898/2026.03.24.26349185

**Authors:** Amsalu Tomas Chuma, Ahmed Saeed Youssef, Melkamu Hunegnaw Asmare, Chunzhuo Wang, Desalew Mekonnen Kassie, Jens-Uwe Voigt, Bart Vanrumste

## Abstract

Reliable interpretation of electrocardiograms (ECGs) requires precise identification of P, QRS, and T (PQRST) wave boundaries. However, it remains challenging due to noise, signal quality variability, and inherent morphological diversity particularly in recordings from children. This study systematically compares the performance of leading deep neural networks (DNN) and heuristic-based delineation algorithms on ambulatory single-lead ECG signals focusing on temporal accuracy. Experiments were conducted using the publicly available LUDB dataset and a private validation dataset comprising 21,759 annotated single-lead wave segments from 611 children recorded using KardiaMobile ECG sensor. DNN were first trained on the LUDB dataset and subsequently tested on the validation dataset. The delineation performance was assessed using Sensitivity (Se) and positive-predictive-value (P^+^) metrics. The best-performing heuristic based and DNN models reached Se and P^+^ of (98.9% vs 97.9%) for P, (99.8% vs 99.2%) for QRS, and (98.7% vs 95.9%) for T wave fiducials, respectively. The lowest standard-deviation (in ms) of wave onset/offset delineation was achieved by attention based 1DU-Net model; ±16.6/±16.3 for P-wave, ±14.0/±16.3 for QRS, and ±26.3/±18.8 for T-wave, respectively. The findings indicate that optimized heuristic models can perform comparably to complex DNN, highlighting their efficiency and suitability for real-time ECG delineation in digital health monitoring applications.

## I. Introduction

The electrocardiogram (ECG) captures the electrical activity of the heart over time during the systole and diastole. The ECG waveform comprises of P wave, QRS complex, T wave (PQRST) and sometimes U-waves defined by an onset, peak and offset fiducial points as illustrated in Fig. 1. A delineation task is to locates these fiducial points in a given ECG. Delineating these fiducials manually is labour intensive and subject to inter-observer variability [1]. The variability arises especially for small or low-amplitude waves or noisy leads. Moreover, delineating offsets for T waves and P-waves during tachycardiac rhythm is ambiguous to decide where its wave plateau ends and baseline returns [1], [2]. This ambiguity leads automatic delineation discrepancies to be bounded under the recommended error tolerance windows of ±150ms according to [2]. And the standard deviation (σ) of the mean delineation error (μ) of each individual wave fiducial point should be smaller than a robust estimate of the error variability expected from evaluations by expert cardiologists, as defined by Common Standards for Quantitative Electrocardiography (CSE) committee [3].

**Fig. 1.**
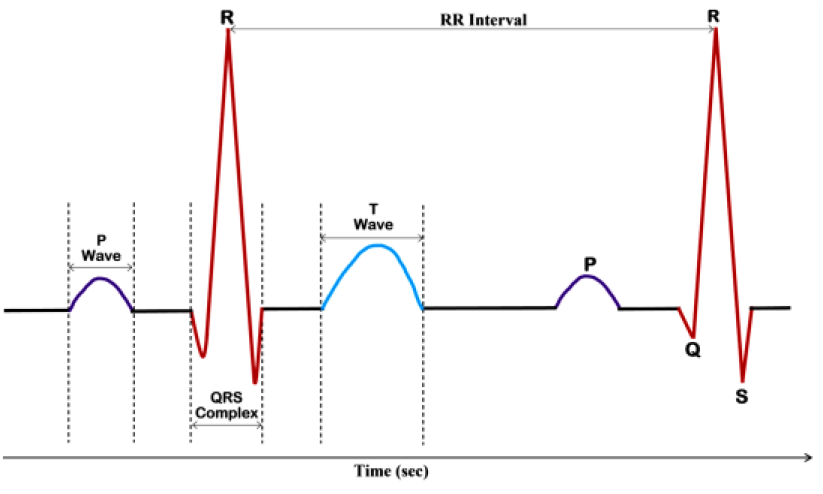
The ECG beat and its key waves. Schematic representation of a single heartbeat from normal ECG showing principal waves (P, QRS, T) and corresponding RR interval

Different automatic delineation algorithms were proposed including rule-based morphology detection [4], wavelet transforms [1], [5]–[8] and template-matching strategies [9], and data driven deep learning frameworks [10]. The wavelet transform (WT) based delineation remains a strong baseline on both single-lead and 12-lead benchmark datasets achieving sensitivities above 97–99% and low σ errors (<16ms) [5], [11]. Heuristic-based delineation methods have been extensively developed and validated, including ECGdeli [5], ECG-kit [6], NeuroKit2 [7], and the more recent visibility graph–based Physio-informed delineator (Prominence method) [12]. Comparative studies have reported strong performance for these approaches, with ECGdeli achieving average sensitivities of approximately 98% for P waves and 92% for T waves on the LUDB and QTDB benchmark datasets [13]. Further evaluations on smartwatch datasets (SmartHeartWatch and SMART Start) showed minimum sensitivities of 92% and 96% for P and T waves, respectively. In particular, the Prominence method has shown to be computationally efficient while achieving performance comparable to deep neural network (DNN)-based approaches on benchmark datasets [12].

Several DNN frameworks have also been proposed for PQRST delineation [10], [14]–[17]. Compared with heuristic approaches, DNN models are computationally demanding and require large training datasets. These models may also suffer from over-segmentation of waveform segments, but offer robust generalization for noisy ECGs. As a result, post-processing techniques are commonly applied to reduce fragmentations and improve delineation accuracy. One widely used approach averages predictions across multiple ECG leads to obtain more reliable waveform boundaries [10], [11], [14]. In addition, modifications to the loss function have been employed to further refine boundary localization [17], [20]. However, these postprocessing and loss-based refinements tend to yield smaller performance gains than those obtained through architectural modifications or more diverse training datasets [10].

In population-screening contexts, particularly in resource-limited settings, wearable or portable devices such as KardiaMobile 6L (KM) provide a practical alternative to standard ECGs. Studies have reported up to 99% agreement for atrial fibrillation detection when KM is compared with standard 12-lead ECGs [21], highlighting their potential for reliable arrhythmia monitoring. Periodic screening with such devices can reduce cardiac morbidity and mortality, particularly in resource-limited, high-prevalence regions. Low-cost automated screening is essential for rheumatic fever and rheumatic heart disease(RHD), which affect millions of children and young adults in LMICs [22]. Evaluating robust PQRST delineation methods on KM recordings from school-aged children is therefore enable automated parameter extraction and facilitating early detection of RHD. Pediatric ECGs have age-dependent morphology, reducing the accuracy of adult-trained algorithms. Compared with standard 12-lead ECGs, this limits spatial information and sensitivity to subtle conduction and repolarization abnormalities.

However, few studies have evaluated KM for schoolchildren and young adults, and none have investigated viability of automated PQRST delineation in ambulatory pediatric and adolescent data. In this study, we used ECG data from the Rheumatic Heart Disease database (RHDdB), collected during a screening campaign in Ethiopia [23] using the KM device, as described in detail in Section II-A. The recordings were evaluated using the best-performing PQRST delineation methods against manually annotated single-lead ECGs, and comparative results are presented. The main contributions of this paper are as follows: first, it validates suitable models tuned for optimal PQRST delineation on KM single-lead ambulatory ECGs. Second, it provides RHD-associated annotated ECGs for external evaluation.

## II. METHODS

### A. Datasets

For the experimental evaluation, two datasets were utilized to develop and assess the PQRST delineation performance comparisons. A publicly available benchmarking dataset, the Lobachevsky University Database (LUDB) [5], was utilized for model training and initial validation. The LUDB containing 58,429 annotated waves of 12-lead ECGs from 200 subjects. In addition, a private ECG recordings from RHDdB was used for external validation of the models.

The RHDdB ECG dataset consists of 30 seconds long Six lead ECGs from 611 children in rural schools of Ethiopia during rheumatic heart disease screening as described in [23]. The data contains 47 RHD positive cases and 564 normal sinus cases based on the confirmatory echocardiography evaluation. The mean ± SD age of the participants is 16.2±2.6 years. The connectivity diagram of KM and the recording smartphone is illustrated in Fig. 2. The procedure for manual PQRST fiducial points annotations, was explained in [24]. The annotations were performed on the lead-II data, and the resultant number of waves annotated for both LUDB and RHDdB is shown in Table I.

**Fig. 2.**
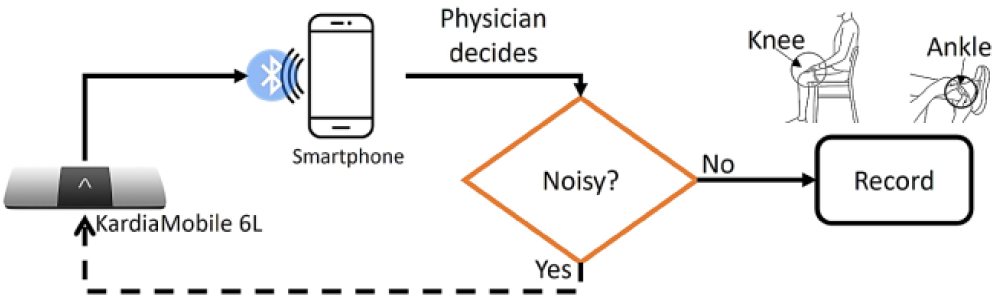
Connectivity diagram of KM to acquire ECG signal. First, the KM device is connected to a smartphone via Bluetooth that has the Kardia app installed. Then, participants place their fingers on the leads according to KM connectivity manual. The recording physician reviews the real-time ECG signal for artifacts, repeating the recording if necessary for subsequent analysis.

**TABLE I.** summary of annotated waves in ludb and rhddb Databases.

| Databases | Leads | P-wave | QRS | T-wave | Total |
| --- | --- | --- | --- | --- | --- |
| LUDB | 12 | 16797 | 21966 | 19666 | 58429 |
| RHDdB | 1 | 7253 | 7253 | 7253 | 21759 |

### B. Preprocessing

The ECG datasets were preprocessed analogous to the steps in [5], [10]. The input ECG waveforms are filtered using a Butterworth zero-phase bandpass filter in [0.5, 100] Hz. An infinite impulse response notch filter was also used to remove powerline noise. The annotations of recordings in LUDB often starts after the first 1-seconds. Hence, the first and last 1-seconds data was cropped out during training. For RHDdB, the annotated 15-second segments were resampled from 300 Hz to 500 Hz to match the sampling frequency of LUDB.

In addition, the training data were augmented following the techniques described in [17]. Transformations included baseline wander, baseline shift, amplitude scaling or resizing, powerline interference, and Gaussian noise. These augmentations resulted in an additional 1,000 trainable 12-lead ECGs derived from the LUDB dataset. No transformations were applied during inference.

### C. Model Selection and Implementation

To perform comparisons on our private RHDdB ECG validation dataset, our evaluation followed the most recent benchmarking models from [10], [12], and the U-Net variants by adding attention gate introduced in [25] at each skip connections. Moreover, publicly available classical and heuristic delineators: ECG-deli [6], Neurokit2 [8], and Prominence method [12] were evaluated. For a given input ECG that has N samples and *d*-leads *x*(*t*) ∈ ℝ^N×*d*^, the model predicts each sample point of a lead, *x*_*i*_, into one of four classes, P, QRS, T or No-wave (isoelectic line).

#### 1) Loss Function

Following the modified hybrid loss function in [17], [20], we trained the remaining DNN models by defining the loss function to enforce the models focus on per sample prediction and contiguity of the predicted classes The categorical cross entropy loss (ℒ_*CCE*_) is calculated for each sample, whereas smoothing constraint (ℒ_*sm*_) adds a penalty when adjacent predictions differ. Analogous to the work of [20], ℒ_*sm*_enforces contiguity of predicted labels in temporal dimension, encouraging adjacent samples to have same class prediction. Hence, the smaller this difference, the smoother the prediction is over time. This can be defined as follows: Let ŷ_i,c_denote the predicted probability of sample *x*_*i*_ belonging to class c, where *i*∈ {1, …, N}indexes the temporal sequence and *c*∈{1,…, C} indexes the classes in an ECG waveform. The ℒ_CCE_ and ℒ_sm_ are defined as:

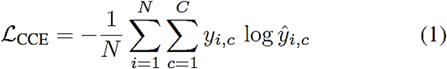

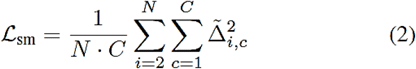

Where the temporal smoothness constraint, 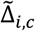, is truncated using a clipping threshold τ, and computed as:

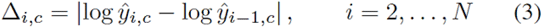

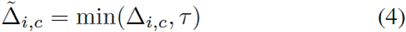

The overall total loss, ℒ, is a weighted sum of LCCE and Lsm:

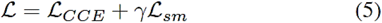

The weight γ for ℒ_*sm*_is initialized as hyperparameters, allowing the model to adjust the balance between classification accuracy and temporal smoothness. Hence, the total loss, ℒ, is a smoothed cross-entropy loss that penalizes the model to avoid overfitting. The other model hyper-parameters tuning procedure was similar to [10]. The hyper-parameter combinations were evaluated based on accuracy on validation set to select the optimal set of hyperparameters. The selected hyperparameters were subsequently used to train the final model.

#### 2) Evaluation Metrics

Following evaluations methods in various similar studies, [1], [5], [10], [12], a detection is considered a true positive (TP) if it falls within a tolerance window (TOL) around a reference manual annotation. Any detected fiducial point with no corresponding manual annotation within the TOL is counted as a false positive (FP), while any fiducial point in the annotation that is absent within the TOL is counted as a false negative (FN). Sensitivity (Se) and positive predictive value (P^+^) are then computed as (Se) = TP/(TP + FN) and (P^+^) = TP/(TP + FP). For prediction errors between the predicted sample point, ŷ, and its corresponding ground truth label, *yi*, within the TOL in ms, the mean error, μ, is calculated as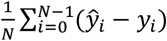, and its standard deviation, 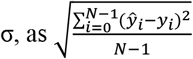 for the detected onsets and offsets of each wave across all records. To correct over-segmentation errors, a simple rule-based method is applied following [10], [17].

Additional evaluations on DNN models by splitting the RHDdB into per-subject stratified five folds while putting the test fold aside and merging the training folds with the training set of the LUDB dataset was reported. This is because of the fact that firstly, the participants in the RHDdB are children and young adults whereas this is not the case in LUDB (16.2±2.6 years vs 52.6±18.1 years). Secondly, the variability in recording device and recorded setting could also attribute to domain bias. Thus, mainly to unbiase the impact of age and device biases on model performance were included in the experiment.

## III. Results

### A. Experiment using Heuristic Methods

The PQRST delineation results for the ECG-deli, Neurokit2 and Prominence methods on RHDdB at standard ±150ms TOL is shown in Table II. The Prominence method achieved best performance over the other open-source delineators reaching Se and P^+^ for all fiducial points above 98%. Relatively lower scores for P-wave with P_on_ (99.13%, 99.24%) while best performed on QRS-wave with QRS_off_ (99.90%, 99.92%), followed by T-wave with T_off_ (98.79%, 99.61%) scores, respectively. Competitive performance was observed with ECG-deli method across all the P and QRS waves, but lower scores for T-wave onsets/offsets, with their corresponding average (Se, P^+^) for P_on_, QRS_off_ and T_off_ of (99.26%, 98.93%), (99.31%, 99.15%) and (96.21%, 95.74%), respectively. The Neurokit method was the least performer particularly for P-wave with about 85.77% Se and large σ error of ±35.7ms.

**TABLE II.** comparison of heuristic pqrst delineation methods on rhdecg dataset at tolerance of 150ms.

| Method | Metric | $P_{on}$ | $P_{off}$ | $QRS_{on}$ | $QRS_{off}$ | $T_{on}$ | $T_{off}$ |
| --- | --- | --- | --- | --- | --- | --- | --- |
| ECG-deli [19] | Se(%) | 99.26 | **99.19** | 99.16 | 99.31 | 95.28 | 96.21 |
| | $P^+$(%) | 98.93 | 98.86 | 99.00 | 99.15 | 94.82 | 95.74 |
| | $\mu \pm \sigma(ms)$ | 24.9±37.3 | -12.8±18.1 | 14.5±14.2 | -25.6±14.2 | -27.4±47.3 | -6.2±46.1 |
| Neurokit [21] | Se(%) | 85.77 | 86.05 | 88.78 | 94.35 | 98.04 | 96.78 |
| | $P^+$(%) | 96.67 | 96.99 | 91.92 | 97.69 | 98.57 | 97.28 |
| | $\mu \pm \sigma(ms)$ | 9.6±35.7 | 50.6±33.1 | 63.5±26.9 | -53.1±33.7 | -35.1±40.9 | 27.5±32.4 |
| Prominence [12] | Se(%) | **99.13** | 98.75 | **99.83** | **99.90** | **98.92** | **98.79** |
| | $P^+$(%) | **99.24** | **99.86** | **99.83** | **99.92** | **99.73** | **99.61** |
| | $\mu \pm \sigma(ms)$ | -10.1±21.6 | -1.7±19.1 | -8.9±11.1 | 6.1±8.8 | -12.6±29.6 | 9.8±19.6 |
| $2\sigma_{CSE}(ms)$ | | 10.2 | 12.7 | 6.5 | 11.6 | N/R | 30.6 |

Importantly, the σ for QRS_*off*_ and T_*off*_ using Prominence algorithm were ±8.8ms, ±19.6ms, respectively. These values are lower than the 2σCSE criteria, indicated in the last row at Table II, for the corresponding upper bound values of the fiducials. The Neurokit was the worst to maintain the σ deviation. As showed in Fig. 3, fiducials delineated for QRS and P waves using Neurokit deviated wider compared to ECGdeli and Prominence. Similarly, as can also be seen in Table II, ECG-deli performed worst for P and T waves fiducial points, particularly P_*on*_ and T_*on*_ points.

**TABLE III.** comparison of deep learning based pqrst delineation methods on rhdecg dataset at tolerance of 150ms.

| Method | Metric | $P_{on}$ | $P_{off}$ | $QRS_{on}$ | $QRS_{off}$ | $T_{on}$ | $T_{off}$ |
| --- | --- | --- | --- | --- | --- | --- | --- |
| 1D U-Net [14] | Se(%) | 95.49 | 97.30 | 96.17 | 97.05 | 85.49 | 90.04 |
| | $P^+$(%) | 91.53 | 93.27 | 95.67 | 96.56 | 76.84 | 94.11 |
| | $\mu \pm \sigma(ms)$ | **-3.8±10.9** | **0.7±10.1** | **6.9±13.1** | **-1.6±15.3** | **-0.5±16.3** | 16.4±22.8 |
| FCN [16] | Se(%) | 97.01 | 97.13 | 98.44 | 98.51 | 95.91 | 96.21 |
| | $P^+$(%) | 96.29 | 96.40 | 99.01 | 99.07 | 94.74 | 95.14 |
| | $\mu \pm \sigma(ms)$ | -12.5±19.0 | -16.1±18.6 | 16.1±15.7 | -0.4±17.1 | -24.1±28.9 | 27.2±28.7 |
| 1D U-Net3+ [18] | Se(%) | 97.65 | 97.69 | 98.53 | 98.56 | 95.61 | 95.44 |
| | $P^+$(%) | 97.44 | 97.48 | 98.65 | 98.71 | 94.72 | 94.32 |
| | $\mu \pm \sigma(ms)$ | -0.5±20.8 | 0.3±18.3 | -6.7±15.4 | 4.0±16.4 | -5.5±28.2 | -9.2±31.2 |
| SegFormer [19] | Se(%) | 92.38 | 93.31 | 96.62 | 95.90 | 90.63 | 89.97 |
| | $P^+$(%) | 91.21 | 91.76 | 97.46 | 97.59 | 91.29 | 90.67 |
| | $\mu \pm \sigma(ms)$ | -14.1±19.6 | -9.7±18.1 | -6.7±22.4 | 9.5±19.3 | -21.3±41.6 | 13.2±31.6 |
| Attention 1D U-Net [25] | Se(%) | **98.30** | **98.37** | **98.87** | **98.92** | **92.96** | **92.82** |
| | $P^+$(%) | **97.42** | **97.49** | **99.20** | **99.25** | **98.77** | **98.63** |
| | $\mu \pm \sigma(ms)$ | -4.8±16.4 | 1.4±16.6 | -8.4±14.0 | 0.7±16.3 | -0.3±26.3 | **15.6±18.8** |
| Attention1D U-Net [25] (5-folds) | Se(%) | 96.15 | 96.11 | 98.57 | 98.08 | 98.86 | 98.59 |
| | $P^+$(%) | 98.04 | 98.08 | 98.42 | 98.59 | 96.65 | 96.39 |
| | $\mu \pm \sigma(ms)$ | -2.0±13.1 | -0.8±12.6 | -4.1±11.6 | 1.4±12.2 | 3.8±22.5 | 6.3±16.7 |
| $2\sigma_{CSE}(ms)$ | | 10.2 | 12.7 | 6.5 | 11.6 | N/R | 30.6 |

The best delineation results are in bold. The five-folds result is shown as a reference. The bottom last row shows the CSE recommended upper-bound limits.

**Fig. 3.**
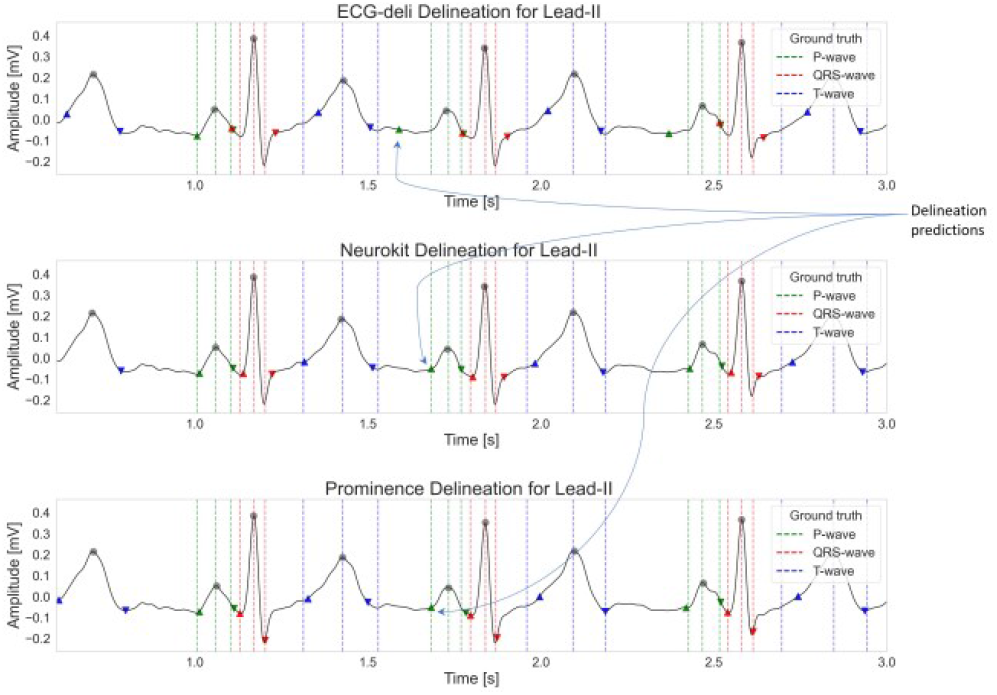
The PQRST delineation result using ECG-deli [6], Neurokit2 [8], and Prominence [12] methods. The dashed vertical lines represent the reference manual annotations of wave onsets/peaks/offsets. The markers indicate delineation algorithms output. Color codes: the P-wave is represented in green colors, QRS in red colors, and T-wave in blue colors.

Following [15], the performance of both heuristic and DNN delineators at more stringent TOLs of 70ms and 40ms are summarized in Table IV. At 70ms tolerance, the Prominence method maintained strong performance, achieving Se and P^+^ above 97%, with σ typically below ±15ms. Under the stricter 40ms criterion, performance decreased modestly, with Se and P^+^ around 93%. Notably, the P_*on*_ and T_*on*_ fiducials exhibited the greatest decline in both Se and P^+^ as TOL decreased, reflecting reduced temporal stability. In contrast, QRS onset and offset demonstrated the most consistent localization (5.7 ± 7.5ms and 9.2 ± 13.2ms, respectively). These results, along with the performance for P_*off*_ and T_*off*_, satisfy the CSE recommended estimates of expected annotation variability by expert cardiologists (see Table IV).

**TABLE IV.**
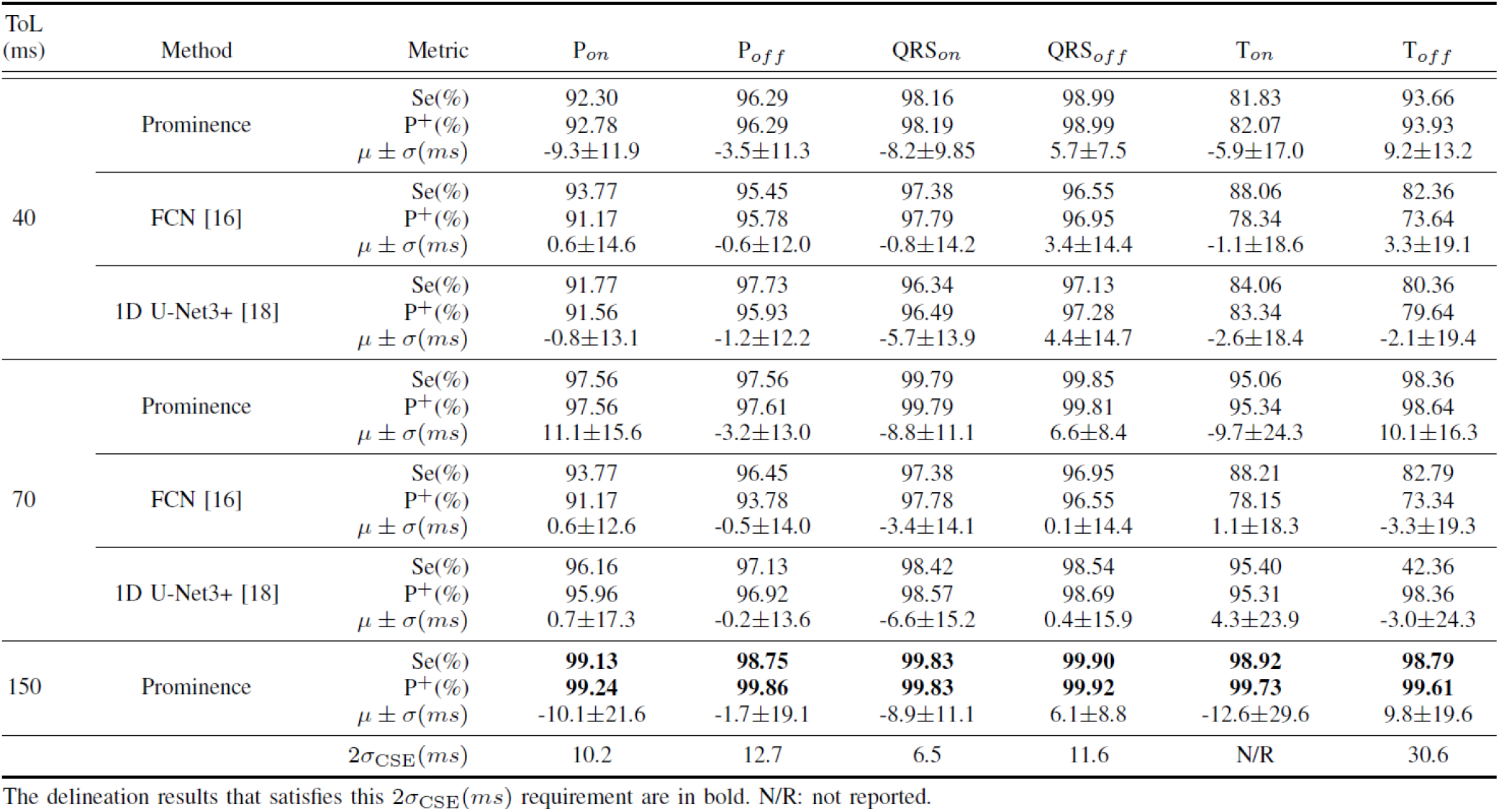
comparison of pqrst delineation result with the manual annotations using the method in [12] on rhdecg dataset at different tolerance thresholds.

Moreover, the results only on 47 confirmed RHD positive cases is shown in Appendix Table V. Similar to the performances on entire RHDdB, the Prominence method showed superior results with the σ for QRS_*off*_ and T_*off*_ of ±10.1ms, ±21.4ms, respectively. At smaller TOL values of 70ms and 40ms, the Se and P^+^ using Prominence algorithm decreased by about 9% for P_*on*_ and 5% for P_*off*_, T_*on*_ and T_*off*_ for each TOL level, while near similar results for QRS fiducials points.

**TABLE V.** Performance of heuristic and DNN methods on RHD positive subjects.

| TOL(ms) | Method |  | Metric | P<sub>on</sub> | P<sub>off</sub> | QRS<sub>on</sub> | QRS<sub>off</sub> | T<sub>on</sub> | T<sub>off</sub> |
| --- | --- | --- | --- | --- | --- | --- | --- | --- | --- |
| 150 | ECG-deli [6] |  | Se(%) | 96.22 | 95.86 | 96.94 | 98.38 | 92.07 | 91.71 |
|  |  |  | P+(%) | 94.35 | 93.99 | 95.73 | 97.15 | 90.60 | 90.25 |
| | | | $\mu \pm \sigma(ms)$ | 22.4±42.1 | -14.9±25.5 | 19.9±18.8 | -23.5±20.0 | -23.5±53.5 | -2.1±56.6 |
|  | Neurokit [8] |  | Se(%) | 79.10 | 72.97 | 79.64 | 79.46 | 98.04 | 96.78 |
|  |  |  | P+(%) | 91.46 | 84.38 | 98.00 | 97.78 | 98.57 | 97.28 |
| | | | $\mu \pm \sigma(ms)$ | 21.8±50.3 | 45.1±53.6 | 11.5±29.9 | -27.1±26.7 | -0.7±31.4 | 8.6±24.3 |
|  | Prominence [12] |  | Se(%) | 96.06 | 93.75 | 98.90 | 99.82 | 97.66 | 98.02 |
|  |  |  | P+(%) | 96.10 | 93.87 | 98.92 | 99.82 | 99.45 | 99.82 |
| | | | $\mu \pm \sigma(ms)$ | -8.1±38.8 | -0.3±32.6 | -1.3±10.8 | 0.7±10.1 | -17.1±31.5 | 1.4±21.4 |
|  | 1D U-Net [14] |  | Se(%) | 97.06 | 97.75 | 98.19 | 98.87 | 92.00 | 92.44 |
|  |  |  | P+(%) | 93.10 | 94.87 | 98.28 | 98.87 | 96.50 | 96.97 |
| | | | $\mu \pm \sigma(ms)$ | -7.1±20.5 | -0.5±21.3 | -3.6±12.9 | -0.4±15.1 | -12.1±33.3 | 11.2±23.0 |
| 70 | Prominence [12] |  | Se(%) | 87.06 | 87.75 | 98.90 | 99.82 | 92.07 | 96.58 |
|  |  |  | P+(%) | 87.10 | 87.87 | 98.92 | 99.82 | 93.76 | 98.30 |
| | | | $\mu \pm \sigma(ms)$ | -14.1±19.1 | -5.8±13.6 | -1.3±10.8 | 0.7±10.1 | -13.1±25.5 | 1.4±17.4 |
|  | 1D U-Net [14] |  | Se(%) | 96.33 | 96.79 | 98.19 | 98.87 | 92.00 | 92.44 |
|  |  |  | P+(%) | 92.92 | 93.36 | 98.19 | 98.87 | 96.50 | 96.97 |
| | | | $\mu \pm \sigma(ms)$ | -7.5±17.5 | -1.7±13.3 | -3.6±12.9 | -0.4±15.9 | -6.1±23.3 | 11.2±17.0 |
| 40 | Prominence [12] |  | Se(%) | 78.06 | 85.95 | 98.90 | 99.46 | 77.84 | 91.89 |
|  |  |  | P+(%) | 79.10 | 85.87 | 98.92 | 99.46 | 79.27 | 93.58 |
| | | | $\mu \pm \sigma(ms)$ | 10.9±12.1 | -5.2±12.9 | 1.1±10.2 | 0.6±9.73 | -8.1±18.5 | 1.6±13.7 |
|  | 1D U-Net [14] |  | Se(%) | 90.06 | 94.75 | 97.05 | 97.73 | 75.78 | 85.78 |
|  |  |  | P+(%) | 87.10 | 91.87 | 97.05 | 97.73 | 79.49 | 89.98 |
| | | | $\mu \pm \sigma(ms)$ | -7.1±10.4 | -2.2±10.6 | -3.1±11.9 | 0.9±15.1 | -3.8±15.5 | 12.3±12.3 |
|  |  |  | 2σ<sub>CSE</sub>(ms) | 10.2 | 12.7 | 6.5 | 11.6 | N/R | 30.6 |

The best delineation results are in bold. The bottom last row shows the CSE recommended upper-bound limits.

### B. Experiment using DNN Models

Comparisons of the DNN models for PQRST delineation performances on RHDdB validation ECG dataset at TOL of 150ms is shown in Table III. The least performed model was SegFormer with Se, P^+^ of (92.38%, 91.21%) for P_on_, (95.90%, 97.59%) for QRS_off_, and (89.97%, 90.67%) for T_off_ waves. The FCN and 1D U-Net3+ models showed better performance than SegFormer model achieving consistent Se and P^+^ of above 96% for P-wave, 98% for QRS, and 93% for T waves. The best delineation results was obtained using Attention 1D U-Net. The Se, P^+^ for P, QRS and T waves reached (98.3%, 97.5%) for P-wave, (98.9%, 99.2%) for QRS-wave, and (92.9%, 98.7%). However, this 1D U-Net model showed low performance for T-wave compared with the other U-Net variant models.

The standard deviation of the delineation error based on 2σCSE(ms) threshold (see bottom last row) indicate the least deviation obtained by 1D U-Net with P_*on,off*_ (10.9, 10.1), QRS_*on,off*_ (13.1, 15.3), and T_*on,off*_ (16.3, 22.8). While the P_*off*_ and T_*off*_ satisfy the CSE threshold, the other fiducials did not. Similar to the separate evaluation on RHD positive cases using heuristic methods in Section III-A, 1D U-Net evaluation yielded an average σ of ±20.8ms, ±13.9ms, and ±27.8ms for P, QRS and T waves, respectively. The results are described in Appendix Table V. None of the obtained σ satisfied the 2σ_CSE_ threshold except for T_*off*_ with ±23.0ms. However, the overall delineation using 1D U-Net are better than the Prominence method particularly for P_*on*_ (-7.1±20.5ms vs - 8.1±38.8ms) and P_*off*_ (-0.5±21.3ms vs -0.3±32.6ms).

Additionally, an overall waveform delineation performance was evaluated using the mIoU metric is showed in Appendix Table VI. At the ±150ms tolerance, the mIoU for P-wave was best achieved using Attention 1D U-Net model with 81.1%. For QRS wave, the Prominence method achieved 83.2% and Attention 1D U-Net for T-wave reaching 81.2%. Considering the overall PQRST waveform delineation, Attention 1D UNet, followed by Prominence showed strong superiority over the other evaluated models.

**TABLE VI.** Performance of heuristic and dnn methods for waveforms using mIOU (%)

| Method Group | Method Name | P | QRS | T | Waveform |
| --- | --- | --- | --- | --- | --- |
| Heuristic | ECG-deli [6] | 67.1 | 67.3 | 60.4 | 64.9 |
|  | Neurokit [8] | 60.6 | 54.8 | 63.7 | 59.7 |
|  | Prominence [12] | **76.7** | **83.2** | **81.2** | **80.4** |
| DNN | FCN [16] | 78.1 | 77.7 | 79.0 | 78.2 |
|  | 1D U-Net3+ [18] | 79.2 | 77.6 | 78.2 | 78.3 |
|  | SegFormer [19] | 64.8 | 70.3 | 58.9 | 64.6 |
|  | Attention 1D U-Net [25] | **80.1** | **81.3** | **81.2** | **80.2** |
|  | Attention 1D U-Net (5-folds) | 83.4 | 84.6 | 83.2 | 83.8 |

## IV. Disccusion

ECG delineation requires precise identification PQRST fiducial points. Our results show that heuristic methods, such as the Prominence method [12], perform competitively with DNN models on single-lead ECGs from KM. Heuristic approaches have linear complexity and require no training data, but depend heavily on ECG quality and fixed search windows, limiting robustness in arrhythmic cases. In contrast, DNNs require large datasets for training and high computational resources.

At a 150ms TOL, both sensitivity and positive predictive value of fiducial markers exceeded an average of 98% for the best-performing Prominence method [12] among heuristic approaches and 1D U-Net [14] among DNN architectures. The σ for these models was confined within 20ms for P and QRS waves, and under 30ms for T-wave fiducials, indicating their suitability for automated delineation of ambulatory single-lead ECG recordings. T-wave boundaries are often prone to subjective bias, leading to larger errors in T_*on*_ and T_*off*_ detection. This arises from the difficulty in identifying the J-point preceding the T-wave onset and the slow transition around its offset, which is often contaminated by noise [1]. As showed in Table IV, these performances were still in acceptable low σ errors at smaller 70ms and 40ms tolerances.

Consistent with previous studies [10], [17], our evaluation showed that encoder-decoder convolutional architectures outperform plain CNNs in detecting waveform primitives, including narrow or inverted QRS complexes and T waves, in challenging ECGs. As shown in Table III and Table IV, U-Net variants demonstrated superior performance compared with transformer-based architectures.

The limitations of transformer-based architectures arise because their attention mechanisms are designed to capture long-range dependencies but have weaker built-in local priors, often requiring larger datasets or stronger pretraining. In the PQRST delineation task, temporal dependencies are largely local within an RR interval, while global context contributes less to identifying fiducial points. Adjustments to patch size, attention dimensions, and the number of heads did not improve performance [10]. Similarly, adding a simple attention gate to the best-performing U-Net yielded only marginal improvement over the baseline model without attention.

Although the Prominence method performed best in this study, heuristic methods generally assume regular rhythm and stable morphology, leading to reduced performance in arrhythmia, ectopy, or low-amplitude waves such as P-waves [1], [12]. Aggregating delineation results across multiple ECG leads can mitigate this limitation. Visual evaluation of six-lead KM data annotated with the Prominence method (Fig. 4) confirms robustness of multi-lead ensembling, shown the last row in the Fig. 4, across different leads. For instance, in the first P-wave of the ensembled output, lower color intensity reflects limited temporal overlap from leads I and aVL. However, such ensemble advantage cannot be done in single-lead analyses. Future work will extend this study to multi-lead evaluations with diverse lead manual annotations for stricter validation of delineation methods.

**Fig. 4.**
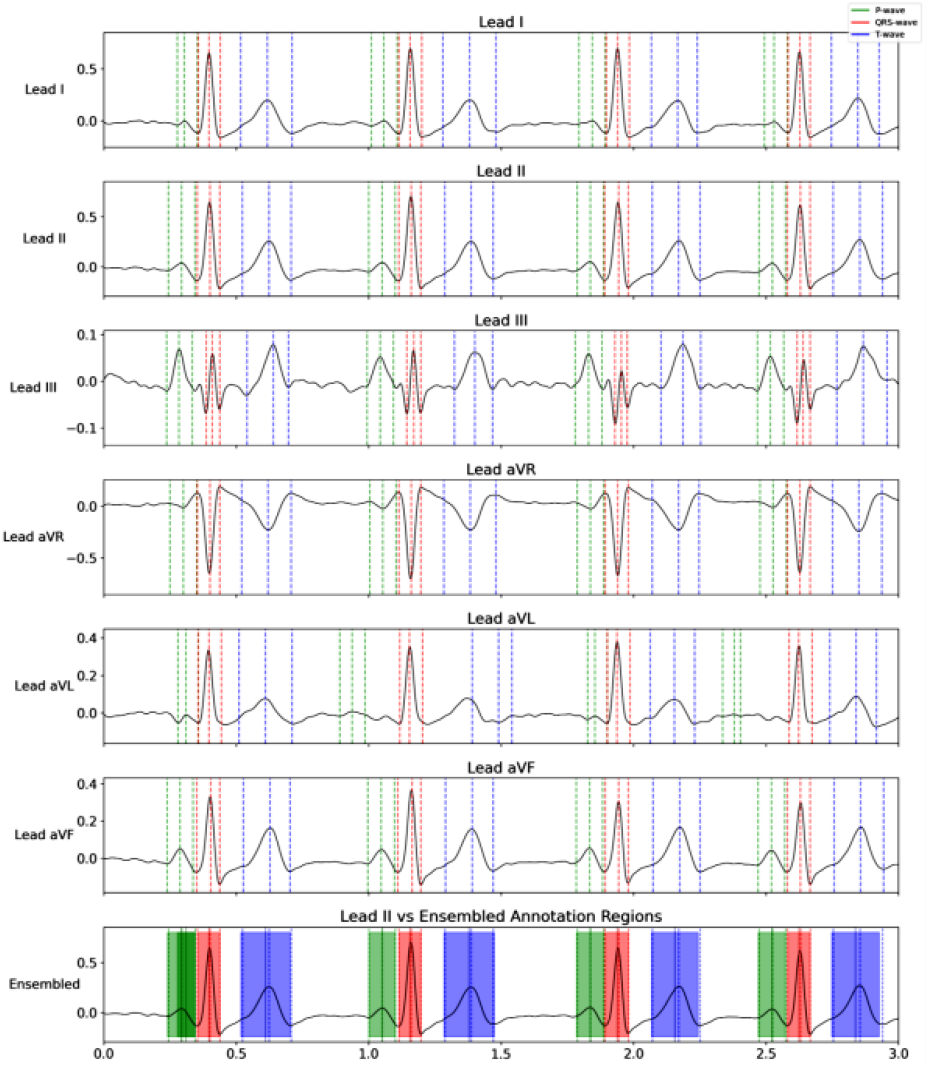
PQRST delineation results using Prominence method [12] on six-leads from KM. The last row indicates the ensemble average of delineation results from individual leads from leads I, II, III, aVR, aVL, and aVF (rows 1 to 6) on lead-II. The amount of temporal overlap is illustrated by color tones, where the lighter the color indicates less overlap. Color codes: green for P, red for QRS, and blue for T waves.

## V. CONCLUSION

Automated delineation of PQRST fiducial points from an ambulatory single-lead ECG was evaluated using different delineation approaches. The performance of heuristic-based Prominence method is competitive to the best DNN model in an overall delineation of single-lead ECG waveform. Importantly, the 1D U-Net variants demonstrated better consistent temporal overlap between automatically detected and reference annotated wave segments than transformers-based and heuristic methods. In resource-limited settings the outperformed methods, particularly Prominence method, can be used as an automated PQRST delineators for single-lead wearable ECG signals. Consequently, this could aid the efforts in automated parameter extraction while screening for RHD cases and early diagnosis at disease prevalent regions.

## Data Availability

All data produced in the present study are available upon reasonable request to the authors.

## ACKNOWLEDGMENT

The study is funded by KU Leuven, center for affordable healthcare with funding reference REF23123123. The authors are grateful to all the participants, study team members and the collaborating hospitals UZ Leuven, TASH and SCH for their unwavering support. We also want to thank flanders AI and Vlaams supercomputer centrum (VSC HPC) for supporting this research. We want to acknowledge the use of OpenAI for editing and grammar enhancement.

## AVAILABILITY OF DATA AND MATERIALS

The RHDECG dataset analyzed in this manuscript is not publicly available. But, can be requested via.

## COMPETING INTERESTS

The researchers have no competing interests to declare.

## APPENDIX

Evaluation of the delineators on RHD positive ECGs (47 subjects) is shown in the Table V. We show the best results from DNN-based 1D U-Net model, and heuristic methods.

Similar to in [10], temporal agreement between predicted and reference ECG intervals was quantified using the mean Intersection over Union (mIoU). For each waveform component, mIoU was defined as the ratio of the temporal intersection to the temporal union of the predicted and annotated segments, averaged across all instances of that component. A higher mIoU values indicate better temporal alignment and delineation accuracy. This metric is independent of the TOL thresholds and can be used to evaluate the performance of delineators on the entire waveform. The Table IV shows the performance of both heuristic and DNN methods for delineating the wave components of the input ECGs.

The Appendix C shows the impact of attention on 1D U-Net with the mIoU metric compared for each wave segments of the input ECG. Although no significant performance gains for overall waveform when attention gate was added, a decreased mIoU for T-wave was observed.

## Notes

### Competing Interest Statement

The authors have declared no competing interest.

### Author Declarations

The study was conducted according to the guidelines of the Declaration of Helsinki and approved by the Ethics Committees of the UZ Leuven with reference No. B3222022001075, Soddo christian hospital No. SCH1941015.

